# Evaluation of nutritional factors, cytokines, liver parameters and survival of patients with liver cirrhosis and hepatocellular carcinoma: a cross-sectional study

**DOI:** 10.1101/2025.01.19.25320796

**Authors:** Iara Carvalho Faria, Leonardo Trevisan Monici, Célia Regina Pavan, Jazon Romilson de Souza Almeida, Sérgio Henrique Dias Marques Faria, Tiago Sevá-Pereira

**Affiliations:** Postgraduate Program in Medical Clinic. University of Campinas (Unicamp), Brazil; Physician. University of Campinas (Unicamp), Brazil; Biomedical. University of Campinas (Unicamp), Brazil; University of São Paulo, Brazil

**Keywords:** Nutritional assessment, liver cirrhosis, hepatocellular carcinoma, cytokines, survival

## Abstract

Hepatocellular carcinoma (HCC) is the leading cause of mortality among cirrhotic patients, often linked to advanced liver disease. This cross-sectional study evaluated the nutritional factors, cytokine profiles, liver function parameters, and survival of patients with liver cirrhosis (LC) and HCC. Forty-seven patients were grouped as LC (n=21) or LC with HCC (n=26). Nutritional status was assessed through anthropometry, bioelectrical impedance analysis, and dietary recall, while cytokine levels (IL-6, IL-10, TNF-α) and biochemical markers (AST, ALT, albumin, prealbumin) were analyzed. Survival data were evaluated using Kaplan-Meier curves and Cox regression. HCC patients exhibited higher IL-6 levels, correlating with advanced disease stages (p=0.035). IL-10 levels were elevated in early-stage HCC (BCLC A) compared to BCLC B (p=0.006). AST and ALT levels were significantly higher in HCC patients, reflecting greater hepatocyte damage. Survival analysis revealed a median of 756 days, with shorter survival in HCC patients (p=0.0172). Nutritional parameters did not significantly correlate with survival outcomes, though most patients were eutrophic or overweight. This study highlights the roles of IL-6 and IL-10 as potential biomarkers in HCC progression and provides critical insights into the biochemical and nutritional profiles associated with LC and HCC. These findings may inform future therapeutic interventions.

## Introduction

Hepatocellular carcinoma (HCC) is a primary cancer of the liver and is considered the principal cause of mortality in cirrhotic patients, with its growing incidence being the sixth most common type of cancer and the fourth leading cause of death from cancer worldwide [1–3]. Patients with liver cirrhosis (LC) are at increased risk of HCC, with more than 80% of these occurring in conjunction with advanced liver disease [4,5].

Liver cirrhosis (LC) is characterized by a functional alteration in the metabolism of macro and micronutrients. The nutritional problems of cirrhotic patients have a multifactorial nature, considering that there is inadequate intake of nutrients caused by anorexia and nausea, in addition to hypermetabolism with poor digestion and absorption [6–9].

Studies have reported the presence of normal weight and obesity in patients with LC [10–12]. Obesity can be an important risk factor for the development of HCC and other serious complications of HCC, such as ascites, hepatic encephalopathy (HE) and infections, contributing to a worsening in quality of life [13,14]. Subsequently, the presence of HCC can induce weight loss due to its association with the synthesis of immunological mediators of the pro-inflammatory response, such as cytokines and chemokines [15].

Cytokines play numerous roles in cell-cell communication at the tissue level, where the outcome is determined by a cytokine concentration and cell type [16]. An important issue related to these signaling molecules is the existence of pleiotropy in the cytokine system. Pleiotropy enables one cytokine to activate several receptors while several others activate the same receptor [15].

Examples of pleiotropic cytokines are tumor necrosis factor alpha (TNF-α), Interleukin 6 (IL-6), and Interleukin 10 (IL-10). TNF-α is a key mediator of the inflammatory response, promoting the release of other cytokines such as interleukin-1 (IL-1) and interleukin-6 (IL-6), and increasing the activity of immune cells such as macrophages and neutrophils [17–19]. TNF-α can induce programmed cell death (apoptosis), particularly in tumor or virus-infected cells. Depending on the cellular environment and the concentration of TNF-α, it can also cause necrosis, a form of cell death that usually causes inflammation in HCC and LC [17–19].

It has been reported that IL-6 is produced by human epidermal cells and various types of tumors [20]. It also plays a key role in inflammation and the innate adaptive system [21,22]. IL-6 stimulates the proliferation of hepatocytes and other liver cells through activation of the JAK/STAT3 pathway, an important pathway for cell growth and survival, which may facilitate the malignant transformation of liver cells [23,24].

IL-10 is produced by macrophages, T-helper2 cells and B lymphocytes and can stimulate and suppress the immune response [20,25]. IL-10 modulates the activity of innate immune cells, such as macrophages and dendritic cells, preventing their excessive activation [25–27]. It also acts on adaptive immunity, affecting the proliferation and function of T and B lymphocytes [26,27]. This contributes to the maintenance of immunological tolerance and prevention of autoimmune responses [26,27].

IL-10 has a dual role in cancer, where it can either inhibit tumor growth by suppressing chronic inflammation or promote local immunosuppression [20,21,28], helping the tumor escape immune surveillance [21,25,29]. In some cancers, elevated IL-10 levels are associated with a worse prognosis, as it may limit the antitumor immune response [30]. Kakumu *et. al.* [31] found that IL-10 levels correlated with serum alanine aminotransferase values, suggesting that this cytokine may also reflect the degree of inflammation in the liver in LC and HCC.

Inflammation caused by increased levels of TNF-α and IL-6, in turn, leads to hyporexia, increases acute phase proteins and reduces negative ones, such as albumin, prealbumin and transferrin [32]. Albumin levels are affected by transcapillary leak, rate of synthesis and catabolism, in addition to external losses from the gastrointestinal tract [33–37]. Albumin synthesis is regulated by hepatic interstitial oncotic pressure while cytokines modulate its gene expression, catabolism and transcapillary [33,38,39]. Hemodynamic stability and normal endothelial function maintain a transcapillary escape of albumin at 4% per hour, which is modified in cases of liver cirrhosis and hepatocellular carcinoma [33].

Due to the ability of carcinoma to alter transcapillary leak, rate of synthesis and catabolism and tissue distribution of albumin, this protein has been extensively used as biochemical markers of survival rate in cancer patients [40–52].

Within this context, the aim of the present study was to evaluate nutritional factors, cytokines, liver parameters and survival of patients with liver cirrhosis and hepatocellular carcinoma. Knowing the characteristics of these patients makes it possible to define future intervention measures.

## Material and methods

This cross-sectional study was carried out in cirrhotic patients treated at the Chronic Liver Disease Outpatient Clinic of Clinical Hospital and the Focal Liver Injury Outpatient Clinic of the Faculty of Medical Sciences, both in State University of Campinas (Unicamp). The study population consists of two groups, one with liver cirrhosis and hepatocellular carcinoma (HCC Group) and the other with liver cirrhosis (LC Group) without neoplasia.

Patients from the HCC group were initially selected, which included individuals with age between > 18 and < 70 years, cirrhotic in Child-Turcotte-Pugh (CTP) [53,54] A or B and HCC according to the Barcelona Classification of Liver Cancer (BCLC) [55, 56] A or B and clinically without ascites with or without the use of diuretics. Subsequently, a similar number of patients were selected for the LC Group with the same inclusion criteria, except for the diagnosis of neoplasia.

Exclusion criteria were: I) history of other malignant neoplasms; II) presence of any degree of hepatic encephalopathy; III) recent gastrointestinal bleeding in the last four months; IV) presence of clinical signs of infection and/or acute or chronic renal failure; V) decompensated diabetes mellitus (glycated hemoglobin: > 6% and/or fasting blood glucose: > 120 mg/dL).

This study was submitted to and approved by the Ethics Committee for Research at the State University of Campinas with the number 1006/2010, and written consent form was obtained from each patient. Personal (date of birth, age, gender) and clinical information (cause of HCC, CTP stage, diagnosis of HCC, BCLC classification) were obtained from the medical record on the day of assessment.

Nutritional assessment was performed after fasting for at least four hours, including anthropometry, bioelectrical impedance analysis (BIA), 24-hour diet recall, cytokine dosage and laboratory measurements. Anthropometry was performed using conventional methods, with a digital platform scale with attached stadiometer (Welmy RIW 200®), clinical adipometer (Cescorf®) and measuring tape (TBW®). Weight, height, mid-arm circumference (MAC) and triceps skinfold thickness (TST) were measured. The following items were calculated: body mass index [BMI = weight/height^2^ (Kg/m^2^)] and the calculation of the mid-arm muscle circumference [MAMC = MAC (cm) – π x TST (cm)]. The anthropometric data collected were compared with the references, being that the BMI data were classified using the World Health Organization (WHO) table [57], while the MAMC and TST were classified using the Frisancho’s table of according to the patient’s gender and age [58].

To verify the body composition, after anthropometry, the tetrapolar bioelectrical impedance test (Maltron Body Composition Analyzer BF906®) was performed with bladder emptying, in the supine position and with prior rest for five minutes. Body fat (BF), basal metabolic rate (BMR), and total body water (TBW) data were obtained. The proportion indicators found in the BIA analysis were classified as low, normal or high according to the equipment references for each patient.

In the 24-hour diet recall, the foods and beverages consumed the day before were reported by the patient in homemade measurements and then entered into a nutrition software (Avanutri® version 4.0) to calculate calories, carbohydrates, lipids, proteins and fibers. The values of macronutrients consumed by the patients were numerically analyzed and the protein content was also quantified in g/Kg of current weight. The obtained results were compared with the recommendations for patients with liver disease (calories and macronutrients) [59] while the comparation to data related fibers were used FAO/WHO tables [60].

On the same day of nutritional assessment, cytokine dosage was performed after a four-hour fasting period, by collecting five mL of blood from the antecubital vein of patients in a disposable syringe and needle and, after to be centrifuged, they were stored in two aliquots of 1.5 mL in micro tubes with lid in a freezer at -20°C. TNF-α, IL-6 and IL-10 concentration analyzes were performed on these samples using the high-sensitivity Enzyme Linked Immuno Sorbent Assay (ELISA) method (manufacturer R&D Systems, INC., Minneapolis, MN, United States) and with the concentrations being expressed in pg/mL.

Laboratory measurements (aspartate aminotransferase (AST), alanine aminotransferase (ALT), albumin, prealbumin and total bilirubin) were measured by the laboratory of Clinical Hospital at Unicamp with collection on the same day or up to three days after the antropometry. The results of these were classified according to the reference standardized by the laboratory.

Patients were followed up for up to 5 years after inclusion in the study to assess survival time. The end date was considered: last follow-up appointment or date of death from any cause recorded in the medical record or system.

Initially, exploratory data analysis was performed using descriptive statistical measures. The assessment of the normality of the distribution of variables was performed using the Shapiro-Wilk test. For comparison of categorical data, the Chi-Square Test was used, and, when necessary, Fisher’s exact test. To compare the non-parametric data between the two groups, Child-Turcotte-Pugh and BCLC, the Mann-Whitney and Kruskal-Wallis test were used and, when necessary, ANOVA. Associations with categorical variables were performed using the Chi-Square Test. For continuous variables, Student’s t test was used. Cox regression analysis was used to assess factors related to overall survival. Survival time analyses were performed using Kaplan-Meier curves. The significance level adopted for this study was 5%, using the Statistical Analysis System (SAS System) for Windows software, version 9.3.

## Results

### Sample Characterization

Initially, 50 patients were selected, however, 3 met the exclusion criteria, 2 due to decompensated diabetes and 1 due to alcoholization of the tumor prior to the evaluation. The sample considered 47 patients, 29 of whom were male (61.7%) and the average age of those evaluated was 54.3±10.8 years, with a minimum age of 22 and a maximum of 69 years.

Separating the data into 2 study groups, named LC Group (n=21), diagnosed with liver cirrhosis and HCC Group (n=26), with liver cirrhosis together with HCC, it can be noted that the prevalence of Child-Pugh A was superior in both groups as well as BCLC A in the HCC group. The HCC group had a higher average age than the LC Group with a statistically significant difference (*p=*0.040). The characterization of the sample and data from the nutritional assessment are shown in Table 1.

**Table 1.**
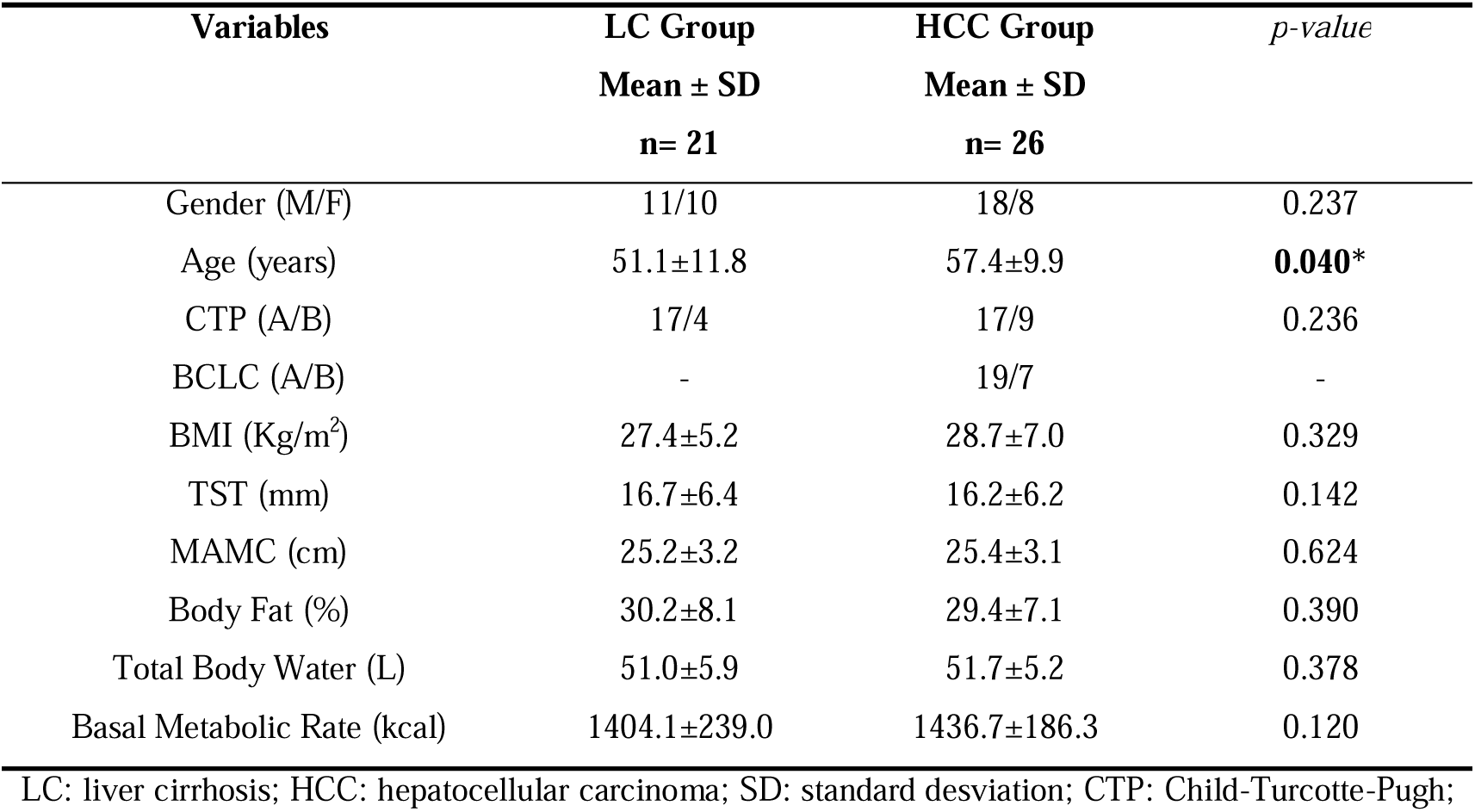
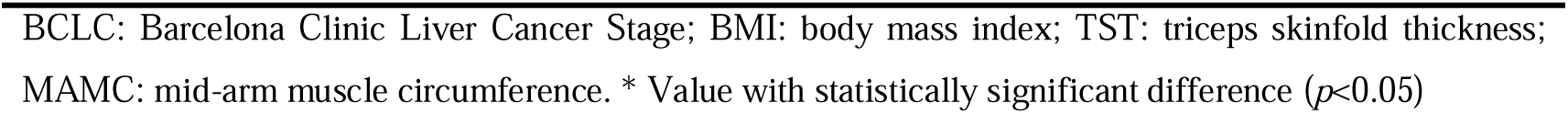
Characterization and nutritional profile of the sample groups.

| Variables | LC Group | HCC Group | *p-value* |
| --- | --- | --- | --- |
| | Mean ± SD<br>n= 21 | Mean ± SD<br>n= 26 | |
| Gender (M/F) | 11/10 | 18/8 | 0.237 |
| Age (years) | $51.1 \pm 11.8$ | $57.4 \pm 9.9$ | **0.040\*** |
| CTP (A/B) | 17/4 | 17/9 | 0.236 |
| BCLC (A/B) | - | 19/7 | - |
| BMI (Kg/m<sup>2</sup>) | $27.4 \pm 5.2$ | $28.7 \pm 7.0$ | 0.329 |
| TST (mm) | $16.7 \pm 6.4$ | $16.2 \pm 6.2$ | 0.142 |
| MAMC (cm) | $25.2 \pm 3.2$ | $25.4 \pm 3.1$ | 0.624 |
| Body Fat (%) | $30.2 \pm 8.1$ | $29.4 \pm 7.1$ | 0.390 |
| Total Body Water (L) | $51.0 \pm 5.9$ | $51.7 \pm 5.2$ | 0.378 |
| Basal Metabolic Rate (kcal) | $1404.1 \pm 239.0$ | $1436.7 \pm 186.3$ | 0.120 |

LC: liver cirrhosis; HCC: hepatocellular carcinoma; SD: standard deviation; CTP: Child-Turcotte-Pugh;
BCLC: Barcelona Clinic Liver Cancer Stage; BMI: body mass index; TST: triceps skinfold thickness; MAMC: mid-arm muscle circumference. \* Value with statistically significant difference ( $p<0.05$ )

Regarding etiology, HCV infection alone was responsible for the cause of liver cirrhosis in 16 (34.0%) patients, followed by alcohol intake with 8 (17.0%) and HBV in 4 (8.5%). The association of HCV and alcohol was responsible for the development of LC in 7 (14.9%) and 1 with HBV+ alcohol. In addition to these etiologies, autoimmune hepatitis was found in 5 (11.4%) patients, primary biliary cirrhosis in 2 (4.3%), non-alcoholic steatohepatitis (NASH) in 1 (2.1%) and cryptogenic in 2 (4.3%).

In the HCC Group, those classified as BCLC stage A (73.1%) and B (26.9%). Of these 26 patients in the HCC Group, 13 patients had a single tumor, while 8 developed two tumors and the other 5 had multiple tumors.

### Nutritional Status and Food Consumption

The data used to assess nutritional status, shown in Table 1, shows that the groups had similar characteristics. The average BMI found in the groups did not show statistically significant difference (*p=*0.329) with values of 27.4±5.2 and 28.7±7.0 Kg/m^2^ in the LC and HCC groups, respectively. The LC Group had patients with normal weight (42.9%), with overweight and obesity having 28.6% each, whereas in the HCC Group, half of the patients were overweight (50.0%), while 30.7% were eutrophic and 19.2% obese.

Assessing the classification of TST and MAMC, most patients in the groups had normality. Body composition by BIA showed no statistically significant difference in patients with LC and HCC. Importantly, total body water was classified as normal in most patients in both groups, while body fat was shown to be high.

Table 2 shows the data on food consumption obtained from the 24-hour recall, and the mean values found for calories, carbohydrates, proteins, lipids and fiber did not show a statistically significant difference between the two groups. The reported caloric intake was higher than the basal metabolic rate analyzed by the BIA in both groups, with 17.1% in the LC Group and 21.2% in the HCC Group. Energy consumption had a mean of 1643.8 calories in the LC Group and 1741.7 calories in the HCC Group.

**Table 2.** Energetic Composition of The Diet Consumption of Groups.

| **Variables** | **LC Group**<br>**Mean ± SD**<br>**n= 21** | **HCC Group**<br>**Mean ± SD**<br>**n= 26** |
| --- | --- | --- |
| Energy (calories/day) | 1643.8±624.1 | 1741.7±677.9 |
| Carbohydrates<br>Requirements: 50-60% of TEV (47) | 59.5±9.6% | 57.9±9.4% |
| Lipids<br>Requirements: 10-20% of TEV (47) | 21.9±6.5% | 23.8±9.2% |
| Proteins<br>Requirements: 20-30% of TEV (47) | 18.6±8.1% | 18.3±6.3% |
| Proteins<br>Requirements: 1.0-1.5 g/Kg/day (47) | 1.1±0.6 g/Kg/day | 1.1±0.6 g/Kg/day |
| Fibers<br>Recommendation: 25-30g/day (48) | 12.8±5.5 g | 17.0±9.7 g |

LC: liver cirrhosis; HCC: hepatocellular carcinoma; TEV: total energy value; g/Kg/day: adjusted to body weight. No statistically significant difference ( $p>0.05$ )

When comparing the dietary intake of macronutrients with the recommended dietary requirements, the LC Group found 47.6% of patients with adequate intake for carbohydrates, 42.9% for lipids and 52.4% for proteins, with the mean protein intake per Kg of current weight was 1.1 ± 0.6g. In this individualized correction, it was observed that most patients in this group had consumption below the recommendation. However, in the HCC Group, 42.3% of patients were observed to have adequate intake for carbohydrates, 42.3% for lipids and 50.0% for proteins, and the mean protein intake per Kg of current weight was identical to the LC Group with the most patients this group (38.5%) also presenting consumption below the recommendation.

### Biochemical Exams

Comparison analyses of cytokine concentrations between each studied group shown in Figure 1 were performed using the nonparametric Mann-Whitney method. In the HCC Group, IL-6 values in BCLC B patients were higher with a statistically significant difference when compared to BCLC A (*p*=0.035). IL-6 concentrations in the BCLC B group were also higher in relation to those belonging to the cirrhosis group (*p*=0.014), however, no significant difference was observed between the values obtained by BCLC A and cirrhosis (*p*=0.145).

**Figure 1.**
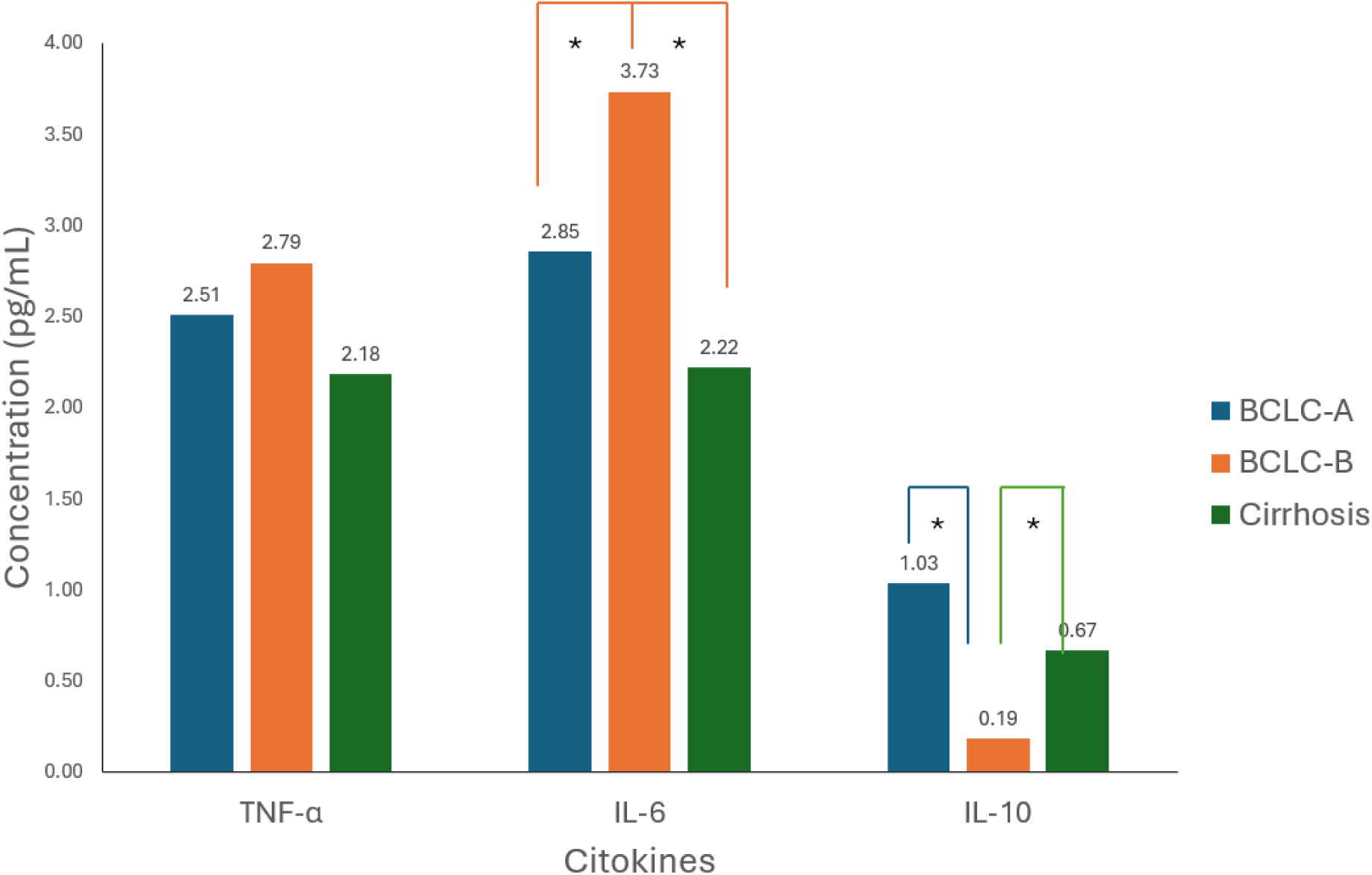
Median concentration of cytokines in pg/mL analyzed in LC and HCC groups (separated by BCLC). LC: liver cirrhosis; HCC: hepatocelular carcinoma; BCLC: Barcelona Clinic Liver Cancer Stage; TNF-α: Tumor Necrosis Factor Alpha; IL-6: Interleukin-6; IL-10: Interleukin-10. * Value with statistically significant difference (*p*<0.05)

Regarding the IL-10 results, it was found a higher median in BCLC A compared to BCLC B, with a statistically significant difference (*p=*0.006) (Figure 1). This situation was also found when comparing the medians of the BCLC B and cirrhosis groups (*p*=0.035). No significant difference was found between the values of the BCLC A and cirrhosis groups, which was different from what was expected, since the size of the difference between the medians suggests the opposite. Considering the TNF-α concentration results, there was no difference between the groups.

The laboratory tests related to liver enzymes and proteins other than bilirubin performed can be seen in Table 3, where significant differences between cirrhotic and hepatocarcinoma patients of AST (*p=*0.037) and ALT (*p=*0.046) were observed by ANOVA test. Comparing the three groups simultaneously, the Kruskal-Wallis test showed a significant difference for the AST variable (*p*=0.043).

**Table 3.**
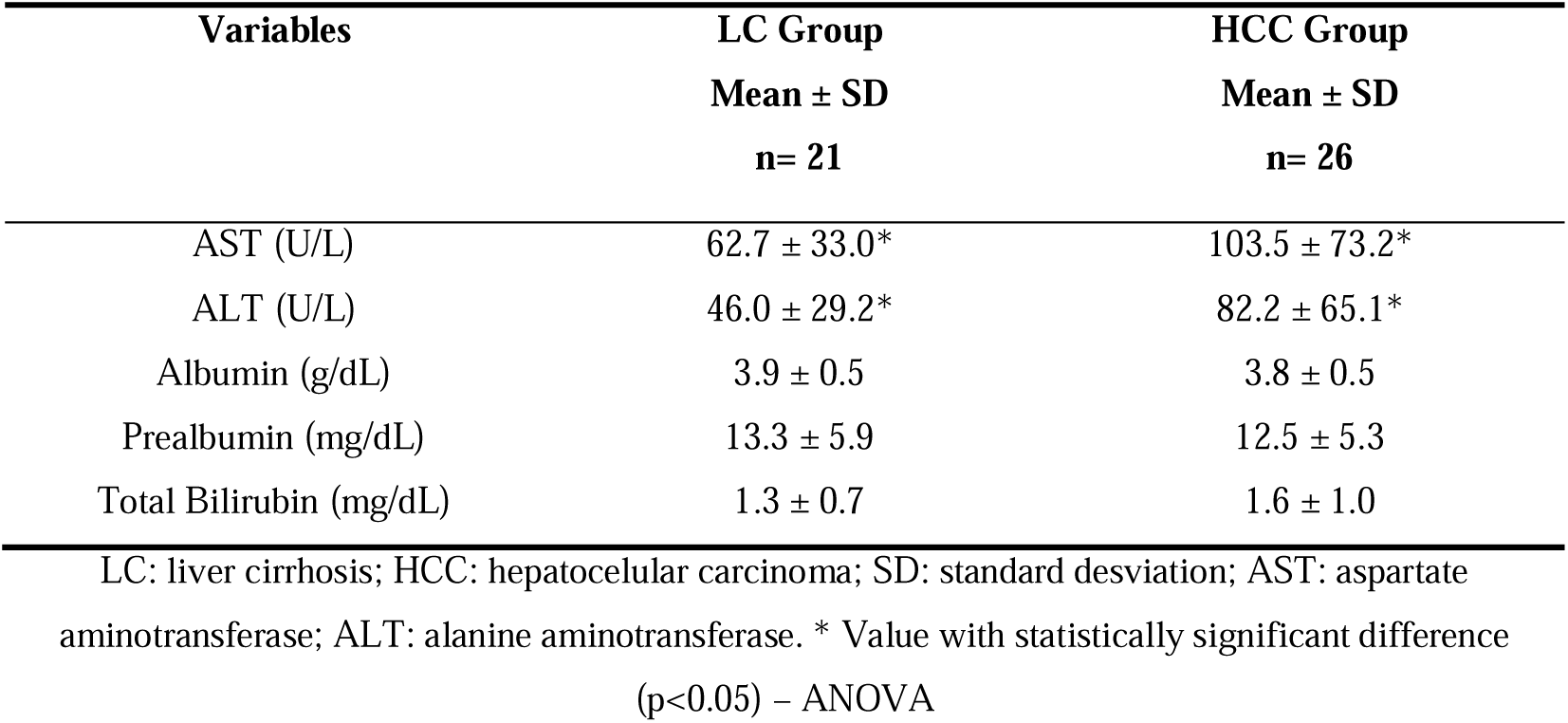
Biochemical tests of study groups.

From the point of view of nutritional markers, the most relevant laboratory tests were albumin and prealbumin, which showed no significant difference between the two study groups (*p=*0.431 and *p=*0.666, respectively).

Analyzing the values found within the HCC Group, separating into BCLC A (n=19) and B (n=7), a statistically significant difference was observed in albumin variable between the values of A (3.9 ± 0.5 g/dL) and B (3.4 ± 0.4 g/dL) with *p=*0.0368. Prealbumin values also show a significant reduction (*p=*0.0184) in the BCLC B (8.3 ±2.0 mg/dL) subgroup compared to BCLC A (13.9 ± 5.3 mg/dL). When checking the levels in relation to the normal value, prealbumin was altered in most of the patients in both groups, with 88.9% being lower in the LC Group and 85.0% in the HCC Group.

### Survival Analysis

Regarding survival time, the median found was 756 days among all patients. Separating cirrhotic patients from those with hepatocarcinoma, the former presented a median survival of 1838 days, while the median survival of patients with neoplasia was 331 days. About the patients who died, 42.9% were from the HCC Group in BCLC A, 28.6% in BCLC B and 28.6% had tumor-free cirrhosis. Uni and multivariate analysis were performed using Cox regression and the data are shown in Table 4. As expected, patients in the LC Group had a longer survival time when compared to those who had a tumor (HCC Group) (*p=* 0.0172).

**Table 4.** Univariate And Multivariate Analysis of Survival Prognostic Factors.

| Variable | Univariate |  | Multivariate |  |
| --- | --- | --- | --- | --- |
|  | HR (CI 95%) | *p* value | HR (CI 95%) | *p* value |
| **Group (LC vs HCC)** | 8.15 (1.45-45.75) | 0.0172* | 164.53 (4.59-5000.0) | 0.0029* |
| **Gender (M vs F)** | 1.03 (0.23-4.63) | 0.9729 | - | - |
| **AST (U/L)** | 1.01 (1.00-1.02) | 0.0216* | - | - |
| **ALT (U/L)** | 1.01 (1.00-1.02) | 0.0929 | - | - |
| **Albumin (g/dL)** | 0.11 (0.02-0.81) | 0.0306* | 12.82 (1.22-142.86) | 0.0099* |
| **Prealbumin (mg/dL)** | 0.82 (0.66-1.03) | 0.0880 | - | - |
| **Total Bilirubin (mg/dL)** | 2.66 (1.06-6.65) | 0.0371* | - | - |
| **Carbohydrate Consumption (in G)** | 1.05 (0.96-1.14) | 0.2862 | 1.26 (1.06-1.51) | 0.0026* |

LC: liver cirrhosis; HCC: hepatocellular carcinoma; HR: Hazard Ratio ; CI: Confidence Interval ; M: masculine; F: feminine; AST: aspartate aminotransferase; ALT: alanine aminotransferase. \* Value with statistically significant difference ( $p<0.05$ ) - Cox regression

The Kaplan-Meier survival curve for cirrhotic and HCC patients did not reach the median (NR), however, there is a significant difference between the two curves (log-rank test *p=*0.007), with a much lower survival for patients with carcinoma (Figure 2). Comparing patients within the HCC Group, BCLC-A against BCLC-B, there was no significant difference in survival time in the Kaplan-Meier curves presenting log-rank test *p=*0.479, although the curves have reached the median. (Figure 3).

**Figure 2:**
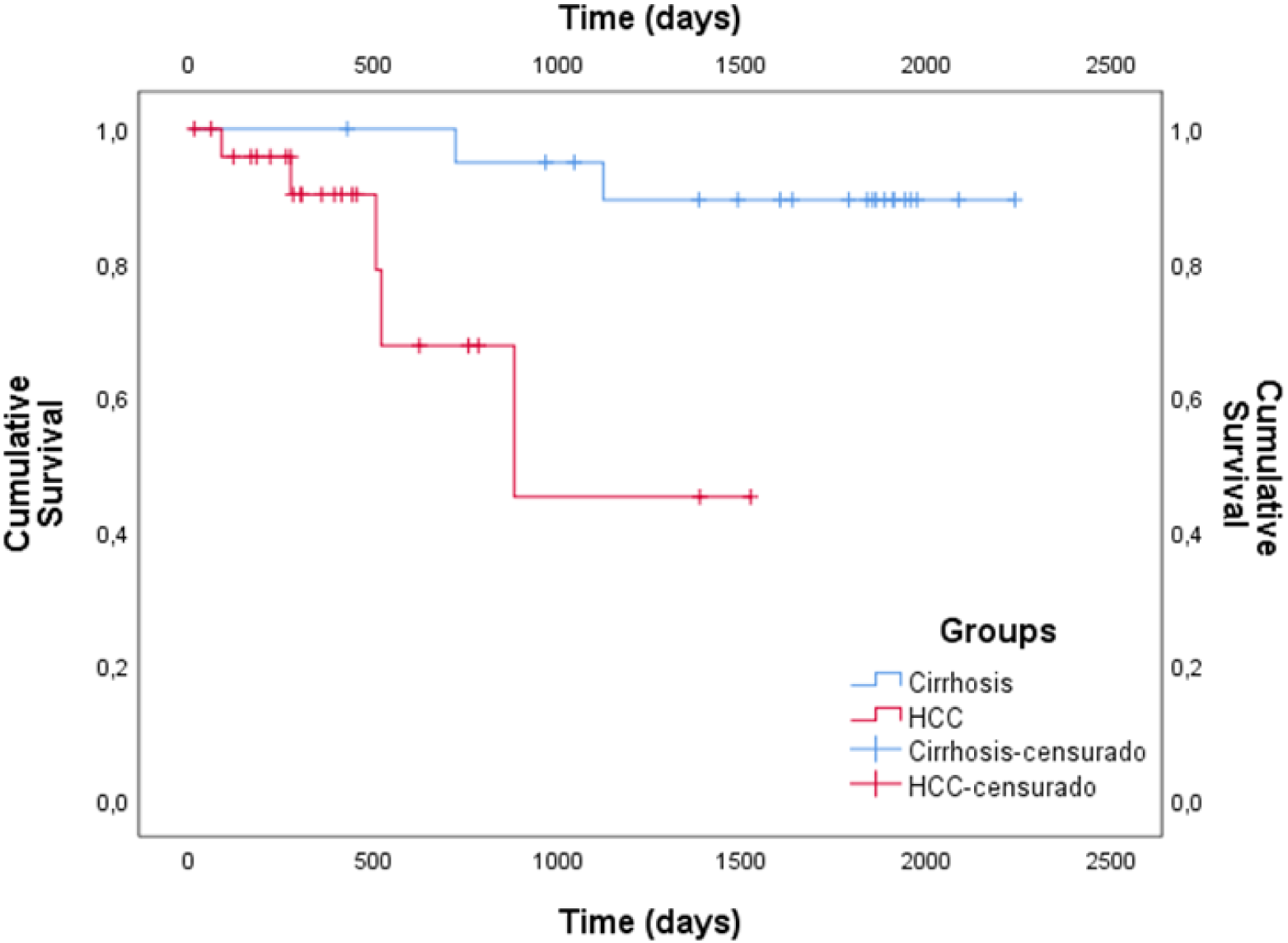
Kaplan-Meier curve shows overall survival in patients with cirrhosis and with hepatocarcinoma (log-rank test, *p=*0.007).

**Figure 3:**
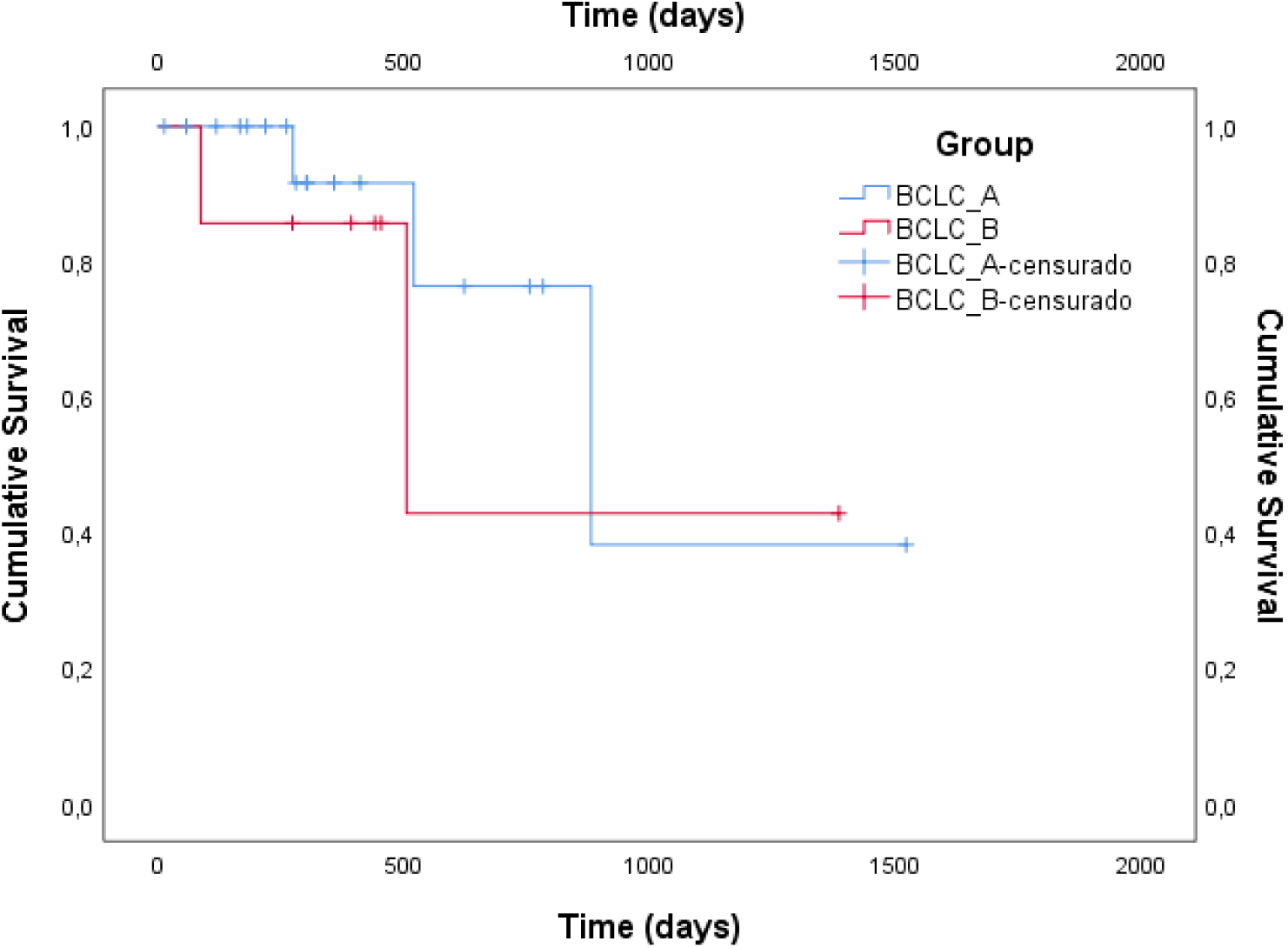
The Kaplan-Meier curve shows overall survival in patients classified as BCLC-A and BCLC-B (log-rank test, p=0.479).

The data in Table 4 from the univariate analysis showed significant differences in survival between the LC and HCC groups for the variables: albumin (*p=*0.0306), total bilirubin (*p=*0.0371) and AST (*p=*0.0261). There was no statistically significant difference in the survival between the groups considering prealbumin.

## Discussion

The etiological and characteristic data of our sample do not differ much from those used in other studies. In the present study, the mean ages were 51.1 and 57.4 years for LC and HCC, respectively, while the mean age for the study of malnutrition in liver cirrhosis by Perez-Reyes *et. al.* [61] was 56.6 years, and in a similar study by Schutte et al it was 58.8 years. Santetti [62] evaluated cirrhotic patients with HCC with cytokine dosage and biochemical tests and found a median age of 60 years, agreeing with Mohamed S. Othman [25] who had a mean age of 55 years in his study and Hsia *et. al.* [30] found patients with a mean age of 59 years.

Analyzing the Child-Turcotte-Pugh score for patients with liver cirrhosis, 81% were in CTP A and 19% in B. Santetti [62] obtained results for cirrhotics of 57.1% in A and 34.3% in CTP B, while Péres-Reyes [61] found in his sample CTP A in 20% and B 57%.

Regarding patients with HCC, our study presented 65.4% in CTP A and 34.6% in B. Van Dijk [63] obtained 82% in A and 16% in B. Schütte *et. al*. [64] used a sample of patients with CTP A of 54.9% and B of 23.5%. As can be seen, both in relation to age and level of liver injury, our sample is very similar to those used in several previously published studies, thus being representative and reliable for making comparisons with the literature of biochemical results and anthropometric evaluation.

Although several studies have reported malnutrition in patients with cirrhosis with or without hepatocellular carcinoma [61,64–67], our study found that most patients in both groups were well-nourished. However, our study is not the first to report patients with chronic liver disease in eutrophy. Van Dijk *et. al.* [63], working on the assessment of the nutritional status of patients with HCC, found that 79% were well-nourished. Gunsar *et. al.* [68], studying the nutritional status of 222 cirrhotic patients, observed 91 well-nourished patients, 58% of whom were in CTP A or B. Maurina [69], on the other hand, nutritionally assessed 208 patients with various types of tumor, 7.7% were obese and 38% were overweight, with 88.8% of the total being well-nourished.

This situation of overweight and obesity found in the sample of this study is in accordance with data from the National Health Survey [70] carried out in 2019, which showed that excess weight affects 60.3% of the Brazilian adult population and that the proportion of obese people in the population with 20 years or over, more than doubled in the country between 2003 and 2019, increasing from 12.2% to 26.8%. This result is well above that estimated by while WHO [71] in which obese people over 18 years were approximately 650 million in 2016 (∼ 13% of the world’s adult population).

The production of cytokines by the body is an inflammatory response that starts with hyperactivity of leukocytes and release of pro-inflammatory cytokines and later produces anti-inflammatory cytokines. This process is involved in liver cirrhosis as well as in hepatocellular carcinoma, probably due to the reduction in liver detoxification ability [32,72].

There is strong evidence reported in the literature on the involvement of cytokines in chronic liver disease such as cirrhosis and hepatocarcinogenesis [25]. One of these cytokines is IL-6, where many studies show its important role in the process of carcinogenesis, because this molecule plays a key role in the process of inflammation and innate adaptive immunity [20,25,73]. In different cases of cancer, its expression is deregulated, increasing its production, making it possible to use its serum concentration as a biomarker for various types of tumors [21,74]. In addition, the increased concentration of this cytokine is correlated with tumor progression [21].

Our data on interleukin 6 in the three groups studied correspond exactly to this scenario. The concentration of IL-6 was higher in patients with more advanced stage of carcinoma, as evidenced by the significantly greater difference in the concentration levels of this cytokine in patients in the BCLC B group compared to those in the BCLC A and cirrhosis (tumor-free) groups. We also found higher concentrations in BCLC A patients compared to patients with cirrhosis alone. This shows that the elevation of IL-6 levels correlates with advanced stages of the disease, in a manner directly proportional to the level of inflammation and liver damage. Our data thus corroborate the possibility and reliability of using IL-6 levels as a biochemical marker of hepatocellular carcinoma.

Regarding IL-10, patients classified as BCLC A presented much higher serum levels of this interleukin when compared to the BCLC B group and the cirrhosis group. Interleukin 10, an anti-inflammatory enzyme with evidence of regulatory action in inflammation-induced fibrosis and carcinogenesis [21], is reported to be at higher levels in patients with HCC [30,75], which is corroborated by our results that indicated a higher amount of this interleukin in the BCLC A group when compared to the other two groups.

The work of Aroucha *et al.* [76] found lower serum levels of IL-10 in patients with high necro-inflammatory activity and greater severity of fibrosis, but with increased levels in moderate and mild liver injuries. It was also found that the higher the TNF-α/IL-10 ratio, the greater the severity of liver injury [76]. This is fully in agreement with our data, where patients in the BCLC B group had a lower level of this cytokine compared to the other groups and consequently a higher TNF-α/IL-10 ratio of 14.7 versus 2.4 and 3.3 in the BCLC A and cirrhosis groups, respectively.

Thus, combining our evidence with that already reported in the literature, it is likely that with the appearance of the tumor, IL-10 levels tend to increase (the increase in IL-10 is directly related to the presence of the tumor itself [75]), since its tumor-induced immunosuppressive activity [21,25,75] acts in the initial development of hepatocellular carcinoma. After the carcinoma is in an advanced stage and has already caused a more severe lesion, there is less secretion of IL-10.

The increased serum levels of aminotransferases in patients with hepatocellular carcinoma are well portrayed in the literature [25,62]. Similarly, our data show that patients with carcinoma have higher levels of AST and ALT when compared with tumor-free cirrhotic patients. Mohammed *et. al.* [25] found a mean of 133.4 u/L and 64.7 u/L of AST and ALT, respectively, while the values of AST of 73.5 u/L and ALT of 60.0 were those obtained by Santetti *et. al.* [62], data very close to those of this study (see Table 3).

The mechanisms for the increased values of these enzymes in the presence of hepatocellular carcinoma are closely related to the degree of hepatocyte necrosis [77]. In patients with hepatocellular carcinoma, there is an elevation of the pro-inflammatory interleukin IL-6 (Figure 1). This cytokine causes chronic inflammation that causes oxidative stress, destroying cell membranes, thus releasing the enzymes AST present in the mitochondria and ALT present in the cytosol of hepatocytes [77]. Another mechanism is tumor invasion and compression in the vascular areas of the liver causing decreased blood flow resulting in the death of hepatocytes [78,79]. Finally, in HCC, cellular regeneration is exacerbated due to the cycle of constant damage and repair of liver tissue. This can result in increased levels of liver enzymes [80]. Even without direct necrosis, cellular proliferation can increase the synthesis and release of ALT and AST [80].

## Conclusions

The sample of this study was very similar to those found in many papers related to the same topic. Unlike several studies that found mainly malnourished patients with HCC, it was observed that the majority of the patients analyzed were eutrophic and overweight, however, there are several reports of the same type found in the literature. The nutritional parameters were shown to be independent factors of the biochemicals analyzed (AST, ALT, IL-6, IL-10, bilirubin, albumin and prealbumin) and no correlation was observed between these parameters and patient survival.

IL-6 levels were shown to be high and directly proportional to the degree of hepatocyte damage, a situation also observed for the aminotransferases AST and ALT. The AST/ALT ratio followed the same logic, being higher in patients with greater liver damage and greater development and progression of carcinoma. Serum levels of IL-10 were higher in patients classified as BCLC A than in those in the BCLC B group. This result indicated a more relevant role for this cytokine in the early development of carcinoma than in states with more developed tumor and greater liver damage.

The Kaplan-Meier survival curves showed greater survival for patients in the LC group (without the presence of tumor) than for those in the HCC group, as expected. However, between the two groups with the presence of tumor, BCLC A and BCLC B, no difference was observed in patient survival. Univariate and multivariate analysis showed an inversely proportional relationship between the concentration of AST and ALT and survival, with patients with low levels of these enzymes living longer. An inversely proportional relationship was observed for the correlation between albumin and patient survival.

Finally, our data demonstrated a detailed profile of biochemical and nutritional parameters and their relationships with the survival of patients with cirrhosis and hepatocellular carcinoma, providing relevant information for the understanding and treatment of these diseases.

## Data Availability

All data produced are available online at Harvard Dataverse

https://dataverse.harvard.edu/dataset.xhtml?persistentId=doi:10.7910/DVN/A81UVO

## Grants and Funding

The author, Iara Faria, thanks the financial assistance (scholarship) from the Coordenação de Aperfeiçoamento de Pessoal de Nível Superior (CAPES).

## Conflict of Interest Statement

None of the authors have any conflict of interest.

## Notes

### Competing Interest Statement

The authors have declared no competing interest.

### Funding Statement

This study was funded by Coordenacao de Aperfeicoamento de Pessoal de Nivel Superior (CAPES).

### Author Declarations

This study was submitted to and approved by the Ethics Committee for Research at the State University of Campinas with the number 1006/2010, and written consent form was obtained from each patient.

